# A Queuing Model for Ventilator Capacity Management during the COVID-19 Pandemic

**DOI:** 10.1101/2021.03.17.21253488

**Authors:** Samantha L. Zimmerman, Alexander R. Rutherford, Alexa van der Waall, Monica Norena, Peter Dodek

## Abstract

We applied a queuing model to inform ventilator capacity planning during the first wave of the COVID-19 epidemic in the province of British Columbia (BC), Canada. The core of our framework is a multiclass Erlang loss model that represents ventilator use by both COVID-19 and non-COVID-19 patients. Input for the model includes COVID-19 case projections, and our analysis incorporates projections with different levels of transmission due to public health measures and social distancing. We incorporated data from the BC Intensive Care Unit Database to calibrate and validate the model. Using discrete event simulation, we projected ventilator access, including when capacity would be reached and how many patients would be unable to access a ventilator. Simulation results were compared with three numerical approximation methods, namely pointwise stationary approximation, modified offered load, and fixed point approximation. Using this comparison, we developed a hybrid optimization approach to efficiently identify required ventilator capacity to meet access targets. Model projections demonstrate that public health measures and social distancing potentially averted up to 50 deaths per day in BC, by ensuring that ventilator capacity was not reached during the first wave of COVID-19. Without these measures, an additional 173 ventilators would have been required to ensure that at least 95% of patients can access a ventilator immediately. Our model provides a tool for policy makers to quantify the interplay between public health measures, necessary critical care resources, and performance indicators for patient access.

## 1 Introduction

The World Health Organization declared COVID-19 a global pandemic on March 11^th^, 2020 [13]. Severe COVID-19 cases may involve critical conditions including respiratory failure, which requires mechanical ventilation for survival [51]. However, hospitals have a limited number of ventilators, which also need to be used by patients with medical and surgical conditions unrelated to COVID-19. The potential surge in intensive care unit (ICU) and ventilator demand due to the pandemic heightens the importance of strategic medical resource management.

The importance of mathematical modeling to project critical care resource demand and capacity requirements during the COVID-19 pandemic is widely recognized [5, 14, 62]. Mathematical models of disease transmission are used to predict COVID-19 case counts [29, 35, 57, 61]. Nevertheless, additional modeling is needed to project medical resource utilization and inform operational decisions [14]. One approach to estimate ICU use is to scale predicted case counts by the projected proportion of cases admitted to the ICU [15, 57]. However, critical care resource utilization also depends on non-COVID-19 demand, resource use time, available capacity, and the delay from COVID-19 symptom onset to critical care. Queuing models can provide an accurate way to project resource utilization by incorporating these stochastic inputs [14].

We applied a multi-class Erlang loss queuing model to inform ventilator management at a provincial level in British Columbia (BC), Canada, during the first wave of COVID-19 in March and April 2020. Our work provides a real-world case study for epidemic capacity planning using Erlang loss models. We used local critical care data from the BC ICU Database to calibrate and validate the model, and incorporated COVID-19 case projections provided by the BC Centre for Disease Control (BC CDC). Model projections capture the interaction between ventilator capacity and patient access. Capacity optimization in the model identifies the number of ventilators required to meet access targets. Model analysis under epidemic scenarios with different transmission levels demonstrates the impact of public health measures and social distancing on ventilator access.

To project ventilator access, we simulated the model using discrete event simulation (DES). We compared three numerical techniques with the simulation results, specifically: pointwise stationary approximation (PSA), modified offered load (MOL) approximation, and fixed point approximation (FPA). To our knowledge, no other studies apply and compare the accuracy of these techniques under the rapid growth of epidemic-type demand, and this represents another contribution of our work.

To inform capacity planning, we identified the number of ventilators required to meet access targets in the model. Loss model performance indicators can capture patient-centred outcomes and limited access to healthcare resources due to capacity constraints. Epidemic-type demand can pose challenges for loss model capacity optimization. Approximations which rely on steady-state formulae may be inaccurate under rapidly changing demand. Furthermore, simulation-based optimization can be especially computationally intensive for queuing models under heavy offered load. To address these challenges, we developed a hybrid capacity optimization approach by combining a simulation-based search procedure with our comparative analysis of PSA, MOL, and FPA. Our hybrid method offers an accurate and computationally efficient approach to epidemic loss model capacity planning. To our knowledge, no other studies use loss model access targets for capacity optimization in epidemic scenarios.

Relevant literature is reviewed in Section 2. The ventilator queuing model is described in Section 3. Model analysis is detailed in Section 4, including access projections and capacity optimization. Section 5 describes BC specific model calibration and epidemic projections, which were used to produce the results presented in Section 6. Lastly, the significance of our results is discussed in Section 7.

## 2 Literature Review

Queuing models play an important role in medical resource and ICU management [7, 11, 12]. The life-threatening conditions faced by many ICU patients motivate the use of Erlang loss models or infinite server models. In loss models, arriving patients are either seen immediately, or are lost to the model if resources are unavailable. In infinite server models, patients are always seen immediately and no capacity limits are incorporated. In both models, patients do not wait for service.

Under non-epidemic conditions, McManus et al. [44] and Julio et al. [36] compared loss model results with historical ICU data and found that they accurately predicted transfer rates. Generalized loss models have been used to determine the required number of ICU beds [42, 52, 63], hospital ward beds [8, 10], and neonatal cots [4]. These studies optimize capacity based on steady-state formulae [4, 10], linear simulation searches [52, 63] or MOL [8]. Although Bekker and de Bruin [8] analyzed of the impact of cyclic patterns in arrival rates, none of these studies consider epidemic growth in arrival rates.

Queuing models can also be used to determine the surge capacity required to meet epidemic or mass causality event demand [38, 41]. Both loss and infinite server queuing models have been used to inform critical care management in response to the COVID-19 pandemic [1, 6, 9, 20, 45, 48, 53, 55, 60]. Infinite server models can capture the stochastic interplay between admissions and length of stay, yielding time-dependent utilization curves that are independent of capacity limits [48]. The probability of excessive utilization can be measured through simulation [20, 53, 55] or exact formulae [6, 9]. However, infinite server models are unable to fully capture performance indicators based on patient access, because these models do not incorporate the impact of finite capacity. On the other hand, loss models work within a finite resource capacity and are able to explore the stochastic relationships between capacity, utilization, and patient-centred access indicators [1, 60]. To our knowledge, loss models have not been used for epidemic capacity optimization.

Time-dependent loss model performance measures can be accurately evaluated using DES. However, this approach is computationally intensive, and a number of numerical approximation methods have been developed [2, 59]. For phase-type service distributions, loss model performance measures can be expressed exactly in terms of the Chapman-Kolmogorov equations [16]. However, solving this system of ordinary differential equations (ODEs) can also be computationally intensive as the system size grows [32]. Approximate ODE solutions include closure approximation [2, 27], Gram-Charlier series expansion [50], and time-dependent perturbation theory[54]. Alternatively, another branch of loss model approximations apply steady-state properties in strategic ways to approximate time-dependent results. In the pointwise stationary approximation (PSA) [21, 23] and its extensions [24, 25], steady-state formulae are applied directly to the time-dependent arrival rate. In the stationary-peakedness approximation, steady-state results are instead applied to an associated non-Poisson model in each time interval [43]. In the modified offered load (MOL) approximation, steady-state results are applied to an offered load given by the expected number of busy servers in an associated infinite server queue [19, 23, 33, 34, 43]. In the fixed point approximation (FPA), time-dependent results and steady-state relationships are applied in an iterative algorithm [3, 32]. Of the above approximation techniques, the three that have existing extensions to general service time distributions and multiple customer classes are PSA, MOL, and FPA. Published numerical comparison of these methods focus on test cases with sinusoidal arrival rates [3, 16, 19, 21, 22, 24, 25, 23, 32, 34]; to our knowledge, no other studies evaluate the accuracy of these methods under epidemic-type growth in demand.

Loss model approximation techniques can be used to identify time-dependent staffing levels required to meet or stabilize loss probability targets, which is often referred to as stabilization. PSA can be applied to determine staffing requirements independently in each time interval, which is termed the stationary independent period by period (SIPP) approach [26, 23]. MOL can be combined with approximate staffing formulas, such as the square root staffing rule [28], to efficiently identify time-dependent staffing requirements [19, 34, 23]. More sophisticated staffing approaches draw on iterative evaluation of performance measures [17, 19, 31]. Li et al. [40] investigate how to achieve stable loss probability during abrupt staffing changes.

## 3 Queuing Model

We modeled pandemic ventilator utilization by applying a two-stage queuing system. The core of this system is a multi-class Erlang loss model, which captures ventilator use by COVID-19 and multiple types of non-COVID-19 patients. Additionally, for COVID-19 patients who require mechanical ventilation, an initial delay model represents the time from symptom onset to the need for ventilation. The complete queuing system for COVID-19 and non-COVID-19 patients is depicted in Figure 1.

**Fig. 1.**
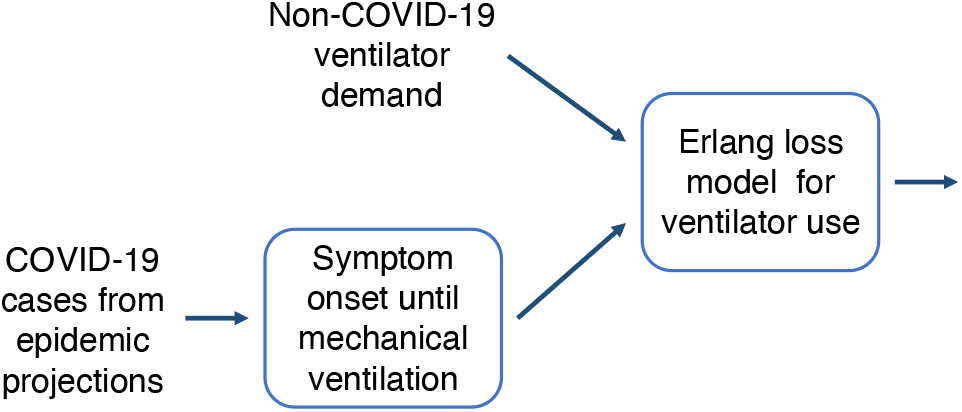
Diagram of a two-stage queuing system for ventilator use by by COVID-19 and non-COVID-19 patients.

The ventilator use model is a multi-class. */G/c/c* Erlang loss model with a limited supply of *c* ventilators that is shared between *K* + 1 groups of patients. It considers both COVID-19 patients (group 1) as well as *K* different types of non-COVID-19 patients (groups 2 through *K* + 1). Each patient requires one ventilator, and if all *c* ventilators are in use, then arriving patients are lost to the system. The use of a loss model for ventilator access is motivated by the life-threatening nature of respiratory failure. Patients who need a mechanical ventilator for life support are not able to wait until one becomes available. When this model is applied to an individual hospital, patients unable to access a ventilator may be transferred to a different hospital. When it is applied at a multi-hospital or provincial level, loss may indicate mortality. Modeling the provincial ventilator supply as a single resource pool assumes that the critical care transfer service is able to work efficiently, even under pandemic demand.

The subdivision of non-COVID-19 demand into *K* groups is based on preliminary data analysis and expert opinion that patient indicators affect the distribution of ventilation time. In our analysis, we considered demand categories based on the diagnoses of viral pneumonia (VP) and acute respiratory distress syndrome (ARDS). Future work could consider other characteristics, such as surgical status, trauma, and age category.

Each of the *K* + 1 patient groups has a potentially different general distribution for time spent on a mechanical ventilator, as well as a different arrival process for ventilator need. We define [*K* + 1] as the set of integers 1 through *K* + 1 representing patient groups. We denote the CDF of each ventilation time distribution as *G*_*k*_, with mean 1*/μ*_*k*_, for all *k* in [*K* + 1]. In the model, non-COVID-19 patients require a ventilator based on non-homogeneous Poisson processes with arrival rate *λ*_*k*_(*t*) at time *t*, for all *k ∈* 2, …, *K* +1. Time-dependency in the arrival rates can capture seasonal patterns in ventilator demand, as well as a reduction in elective surgeries in response to COVID-19. For COVID-19 patients needing mechanical ventilation, the effective ventilator demand is the output of the symptom delay model.

Epidemic case projections are translated into ventilator demand using an *M*_*t*_*/G/∞* queuing model to represent symptom delay. This infinite server model does not correspond to utilization of any physical resource; it simply implements a stochastic delay from the onset of COVID-19 to the presentation of severe symptoms requiring mechanical ventilation. The arrival process for this model is a non-homogeneous Poisson process with rate *λ*_0_(*t*) at time *t*, which we base on scaled localized COVID-19 case projections. The output process of the symptom delay model is a non-homogeneous Poisson process, with rate *λ*_1_ given by the convolution [18],

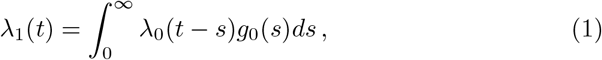

where *g*_0_ is the symptom delay PDF. In other words, ventilator demand is a weighted average of past case rates, scaled by the proportion of critical cases and weighted by the symptom delay PDF.

A key system performance measure is the loss probability at any time *t*, denoted by *β*_*c*_(*t*). This represents the probability of a patient being unable access mechanical ventilation, given that they need a ventilator at time *t*. Other system quantities of interest include the time-dependent expected number of ventilators in use and offered load, denoted by *m*(*t*) and *a*(*t*) respectively. The patient group specific version of these quantities are denoted by *m*_*k*_(*t*) and *a*_*k*_(*t*), for all *k* in [*K*+1]; however, the loss probability *β*_*c*_(*t*) is the same for each patient group. To our knowledge, there are no exact analytical formulas for *β*_*c*_(*t*), *m*_*c*_(*t*), and *a*_*c*_(*t*).

## 4 Model Analysis

Our queuing model analysis focuses on addressing two practical ventilator management questions. Firstly, we projected ventilator access in the model based on epidemic forecasts. These projections determine whether the ventilator supply is sufficient, when capacity would be reached, and the rate of patients unable to access a ventilator. Comparing projections under multiple epidemic scenarios can link public health measures to ventilator access. Subsection 4.1 describes the four methods that we applied and compared for projecting ventilator access, namely: discrete event simulation (DES), pointwise stationary approximation (PSA), modified offered load approximation (MOL) and fixed point approximation (FPA). Secondly, we optimized ventilator capacity by identifying the minimum number of ventilators required to keep the loss probability under 5%, over the course of a projected epidemic scenario. This optimization problem identifies a single peak requirement, since this is sufficient to inform planning. Unlike staffing, which can fluctuate to address demand, ventilator capacity does not vary day to day. Subsection 4.2 describes our hybrid approach to capacity optimization that draws on the numerical approximations in Subsection 4.1.

### 4.1 Projecting Ventilator Access

We applied and compared four methods for approximating ventilator access in the model. DES is an accurate estimator of time-dependent system performance in queuing models and it is often used for analysis or bench marking of intractable models. However, simulation can be computationally intensive, especially under heavy offered load. On the other hand, the numerical approximations PSA, MOL, and FPA are all computationally efficient, albeit with less accuracy. PSA is typically inaccurate when arrival rates change rapidly relative to service times [21, 22]. Although MOL is often more accurate than PSA, it has declining accuracy for higher loss probabilities [43]. FPA has demonstrated high accuracy in published test cases [3, 32]. Previous numerical comparisons of these methods have solely used sinusoidal test cases [3, 16, 19, 21, 22, 24, 25, 23, 32, 34] and not epidemic arrival rate functions. Subsubsection describes our DES implementation, which we use as a benchmark for three other numerical procedures. Our applications of PSA, MOL, and FPA are described in Subsubsections 4.1.2, 4.1.3, and 4.1.4, respectively.

#### 4.1.1 Discrete event simulation

We built a DES simulation of the queuing model using the AnyLogic modeling software. The simulation has two source nodes, one for COVID-19 and one for combined non-COVID-19 patient streams. Each source node generates patients according to separate non-homogeneous Poisson processes, which are each obtained by thinning a homogeneous Poisson process with a time-dependent probability of acceptance. Generated non-COVID-19 patients are then randomly assigned into groups based on time-dependent mixing probabilities. Generated COVID-19 patients face an additional stochastic symptom delay prior to ventilator use. A single service block combines ventilator use for all patient types. Simulated patients will use any available ventilator, with a service time that is generated randomly from a distribution based on patient group. If all ventilators are in use, then arriving patients will leave the model without returning or affecting future ventilator use.

At regular time intervals, the DES model measures the number of ventilators being used and whether capacity has been reached. Averaging these measurements across multiple simulation runs yields time-dependent estimates 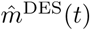 and 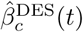 for the expected ventilator use and loss probability, respectively, at each sampled time *t*. Confidence intervals and interquartile ranges were also calculated for these estimates.

#### 4.1.2 Pointwise stationary approximation and steady-state formulae

The PSA approach obtains proximate expressions for time-dependent properties by substituting the time-dependent arrival rate into steady-state formulae [2, 21] at each time point. PSA assumes that the system reaches a new equilibrium value instantaneously as the arrival rate changes, and does not incorporate the impact of past arrivals. Some extensions try to mitigate this by incorporating a lag in arrival rate substitution [24, 25]. While this approximation is computationally efficient, it is known to be over-responsive and have limited accuracy in scenarios with arrival rates that change rapidly relative to service times [21, 22, 23]. We applied PSA by substituting estimated ventilator arrival rates into steady-state formulae for our multi-class loss model.

If the arrival rate were time-independent, then the loss model would have an equilibrium state with constant loss probability. Even though ventilation time is generally distributed, Takács [56] proved that general service time loss models have the same steady-state distribution as exponential service time loss models, under time-independent arrivals. Furthermore, Kaufman [37] proved that this property of insensitivity to service time distribution also extends to multi-class loss models. The steady-state loss probability, *β* is given by Erlang’s B formula,

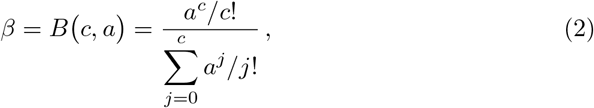

where *c* is the number of ventilators and *a* is the constant total offered load. Erlang’s B formula is an increasing function of *a* and a decreasing function of *c*. For ease of calculation, *B*(*c, a*) has an equivalent recursive expression [46]

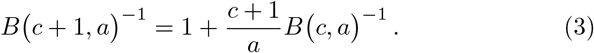

The total offered load *a* is given by the sum of the constant offered load for each group,

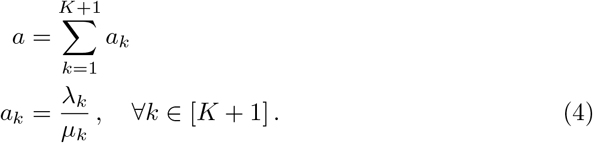

The steady-state expected number of busy ventilators for patient group *k* is given by

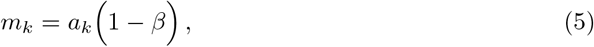

and the total over patient groups is

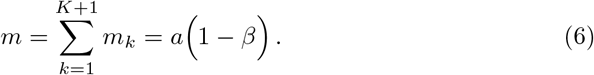

Applying equation (2), the PSA proximate loss probability 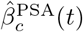 of the ventilator model, with PSA time-dependent offered load 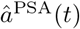, is given by

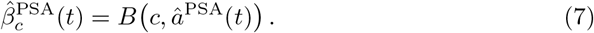

Furthermore, the PSA estimate for the expected number of busy servers is

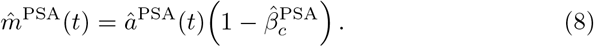

In a multi-class loss model, the PSA time-dependent offered load is not dependent on *c*, and given by

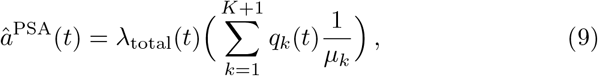

where the total arrival rate is

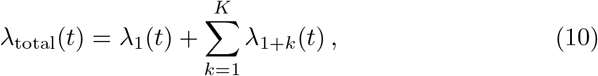

and the average service time is weighted by the instantaneous proportion of arrival rate for each group *k* in

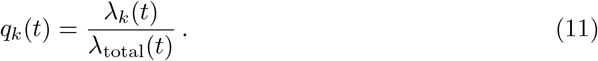

#### 4.1.3 Modified offered load

In the MOL approach, time-dependent offered load is approximated by utilization in a corresponding system without capacity limits [19, 33, 34, 43]. In this infinite server system, the expected number of busy servers, denoted by *m*^*∞*^(*t*), incorporates the interplay between past arrivals and service times [16]. Substituting this offered load estimate into steady-state formulae provides an approximation for finite system performance measures.

The MOL estimates for offered load, loss probability, and the expected number of ventilators in use are

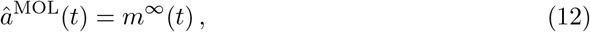

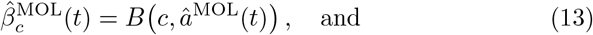

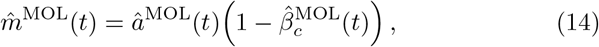

respectively. Erlang’s B formula *B* (*c, a*) is given by equations (2) and (3). Here *m*^*∞*^(*t*) is a sum over patient groups,

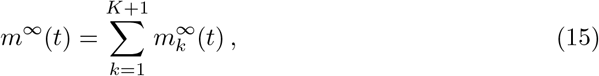

where 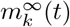 is the expected number of ventilators in use by patients of group *k* in an infinite server system. Without capacity limits, there is no patient interaction, thus each 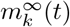 is individually given by the convolution [18],

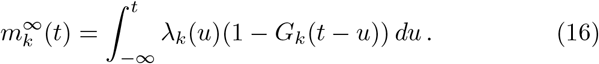

#### 4.1.4 Fixed point approximation

The FPA approach iteratively estimates performance measures at several fixed points in time [3, 32]. Iterative estimates of the time-series for loss probability, utilization, and offered load are sequentially updated, until consecutive changes are insubstantial. Each iteration refines estimates of these performance measures using both steady-state and time-dependent relationships. By starting with an initial loss probability of zero, the FPA algorithm begins with MOL values and iteratively improves estimates to incorporate the impact of finite server capacity [3, 32].

We applied the multi-class FPA algorithm described by Izady and Worthington [32] to our loss model. For each iteration *i*, the loss estimate is denoted 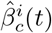. The corresponding estimate for the expected number of ventilators in use is given by

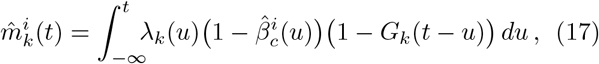

for all *k* in [*K* + 1]. Iterative offered load is estimated using the steady-state relationship in equation (5), rearranged to give

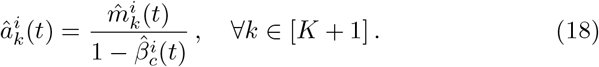

The sum of offered load estimates is substituted into Erlang’s B formula (2) or (3), to yield a subsequent estimate of loss probability,

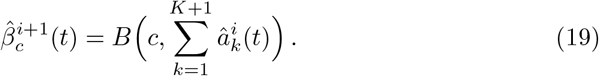

The FPA algorithm repeats the three steps given by equations (17), (18), and (19) in each iteration, until sequential loss probability estimates are within a set tolerance of each other. The FPA approach uses regularly spaced time points and numerical integration in equation (17) must be based on fixed and regular time points.

### 4.2 Optimizing Ventilator Capacity

To address ventilator capacity planning, we determined the minimum number of ventilators required to maintain modeled loss probability under target *α* over the time planning horizon *T*, which ensures 100(1 *− α*)% access. This capacity optimization problem can be formulated as

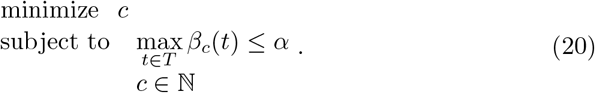

Maximum modeled loss probability is a strictly decreasing and non-linear function of the number of ventilators *c*. This response function can be approximated by the methods described in Subsection 4.1, namely DES, PSA, MOL, and FPA, which give the estimates 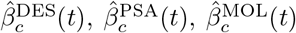, and 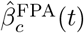), respectively. Substituting each of these estimates into problem (20) provides proximate capacity requirements.

Simulation is the most accurate of these methods to estimate the response function; however, solving the optimization problem (20) using DES can be computationally intensive. On the other hand, the three approximation methods PSA, MOL, and FPA are computationally efficient, but may be inaccurate. We compared three proximate capacity requirements—obtained by substituting PSA, MOL, and FPA estimates into problem (20)—with the DES solution. To boost the efficiency of our DES search, we developed a hybrid optimization approach with a strategic starting point informed by our comparison of approximation methods to project access under the current ventilator supply.

To approximately solve problem (20) using PSA, MOL, and FPA, we simply performed deterministic linear searches, by increasing the ventilator capacity until each loss estimate 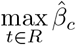 is under *α*. This straightforward approach sufficed because of the computational efficiency of these approximation methods.

To solve the optimization problem (20) using DES estimation for the response function, we applied a modified response surface methodology (RSM) search procedure to incorporate the stochasticity in 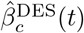. Our response function is only defined on natural number capacities; consequently, multi-point root-finding procedures are more suitable than single-point methods, which rely on derivative proxies [49]. The RSM framework typically uses first and second-order approximations of a sampled response function to guide an optimization search [47, 39]. We modified the RSM framework in Nicolai and Dekker [47] for stochastic root finding. Our adapted procedure is guided by second-order approximations of the response function, to account for non-linear and asymptotic behavior as capacity grows. In each iteration, the algorithm identifies a root with *α* of a second order approximation. This intersection point is used as a subsequent solution estimate, which becomes the centre-point for re-approximating the response function. The algorithm is repeated until desired convergence in centre-points is achieved.

In our application, we determined an initial centre-point for RSM using a proximate required capacity, obtained from the numerical approximation method which best projected current capacity outcomes. We ended the algorithm at a change of 5 ventilators, and plotted the loss probability for the ten capacity values surrounding the final centre-point. A graphical representation enables further interpretation of our results by decision makers, and illustrates the sensitivity of loss probability to the number of ventilators. Appendix A discusses the algorithmic details of our modified RSM and linear search approach.

## 5 Data Analysis and Case Projections

We analyzed data on critical care utilization in BC, in order to apply our model to the BC context. The primary data set used was an extract from the BC ICU Database. We supplemented this with summary data provided by the BC Ministry of Health from the Discharge Abstract Database, published reports, expert opinion, and data from the Provincial Health Services Authority on ventilator capacity. The BC Centre for Disease Control provided case projections for the COVID-19 epidemic in BC as input for the model.

The British Columbia ICU Database was established in 1998 at the Centre for Health Evaluation & Outcome Sciences to provide detailed information on the delivery of critical care in British Columbia [58]. Our data extract consists of records from calendar years 2016–2018. At the time of this work, 2019 data was unavailable. For 2016–2018, the database contains ICU data from 20 hospitals in BC, including nearly all major hospitals. However, an additional 21 hospitals in BC with ICUs are not included in our extract. For these hospitals, we used ICU admission data from the Discharge Abstract Database, which is a national database of hospital admissions in Canada. We estimated ventilator utilization for these hospitals by assuming that they have the same fraction of ICU admissions requiring mechanical ventilation as the hospitals in the ICU Database. Our ICU Database extract includes an entry for each instance of mechanical ventilation. It has fields for the start and stop time of mechanical ventilation, acute respiratory distress syndrome (ARDS) diagnosis, and viral pneumonia (VP) diagnosis. We included these diagnoses in our extract because they are clinically similar to respiratory failure due to COVID-19.

### 5.1 Non-COVID-19 Demand and Ventilator Capacity

We projected non-COVID-19 ventilator demand into 2019–2020 by using the frequency of ventilation starts in the most recent two years of our BC ICU Database extract, namely 2017–2018. Patients may have multiple ventilation periods during a single ICU stay; however, we treated these as independent ventilation starts. Figure 2 shows a monthly time series of mechanical ventilation starts in the BC ICU Database for 2017 and 2018. For both years, the rate of ventilation starts is higher at the beginning of the year than at the end; however, 2018 has an additional peak in May. To balance capturing seasonal tends without over-fitting to these years of data, we projected ventilation demand into 2019–2020 using the average monthly rate over both years, which is also shown in Figure 2. We multiplied this by 1.198, which was computed from the DAD to account for the additional 21 hospitals not in the BC ICU Database.

**Fig. 2.**
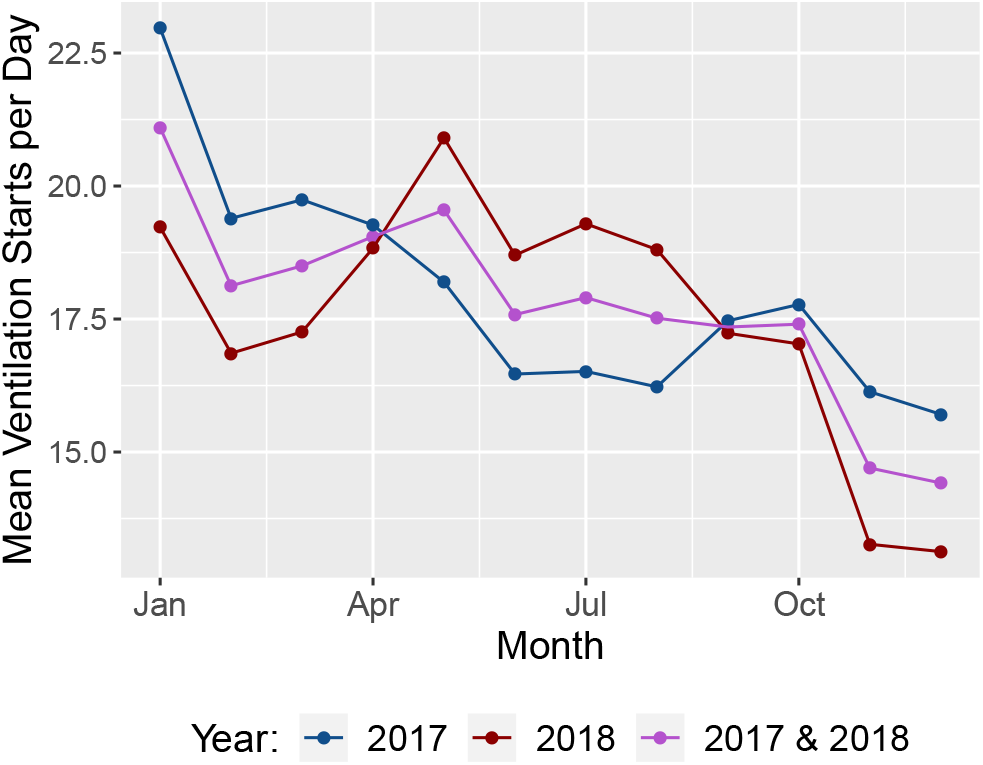
Average monthly rate of mechanical ventilation starts from the ICU Database, for years 2017 and 2018, along with a combined monthly average across both years.

The BC Ministry of Health responded to the COVID-19 pandemic by canceling non-urgent elective surgeries as of March 16^th^, 2020. Detailed data on the impact of this reduction was unavailable at the time of this study. Based on expert opinion, we estimated that this led to a 15% reduction in the number of non-COVID-19 patients requiring mechanical ventilation. We implemented this change as a step reduction the non-COVID-19 demand rate by 15% from March 16^th^, 2020 onwards.

The Provincial Health Services Authority of BC conducted an inventory of ventilators in the province in March 2020. There were 498 adult mechanical ventilators available in 34 hospitals. We were advised that at any given time, approximately 10% of these ventilators would be unavailable due to repair or maintenance. Therefore, we set the current number of ventilators in the model to 448. Based on consultation with a respiratory therapist, we assumed that the time required to clean a ventilator and prepare it for a new patient is approximately two hours, which we incorporated into the modeled ventilation service time.

### 5.2 Mechanical Ventilation Time and Symptom Delay

We characterized the duration of time that patients receive ventilation by using start and stop times from the 2017–2018 records of our BC ICU Database extract. Approximately 10.4% of these records were either missing a start/stop time or had zero ventilation time, and we did not use these records in ventilation time analysis. We divided the remaining records into two groups: one for patients diagnosed with either ARDS or VP, and one for patients with neither of these diagnoses. At the time of this analysis, there was limited data for ventilation time of COVID-19 patients. ARDS and VP are clinically similar to respiratory failure due to COVID-19. Therefore, we assumed that the distribution of ventilation time for patents with these diagnoses is representative of the distribution for COVID-19 patients.

For each group of records, we fit ventilation time to a gamma distribution using the maximum-likelihood method implemented in the ‘MASS’ package in R. The parameters for the distribution fits are given in Table 1 and the distributions are plotted in Figure 3. The ventilation time data appears to be well captured by the fitted gamma distributions. The mean ventilation time for patients with VP or ARDS is substantially greater than the mean ventilation time for patients with neither diagnosis.

**Table 1.** Fitted gamma distribution parameters for time spent on mechanical ventilation.

|  | VP or ARDS | Neither VP nor ARDS |
| --- | --- | --- |
| Shape | 0.94 | 0.85 |
| Scale | 7.9 days | 4.8 days |
| Mean | 7.5 days | 4.1 days |

**Fig. 3.**
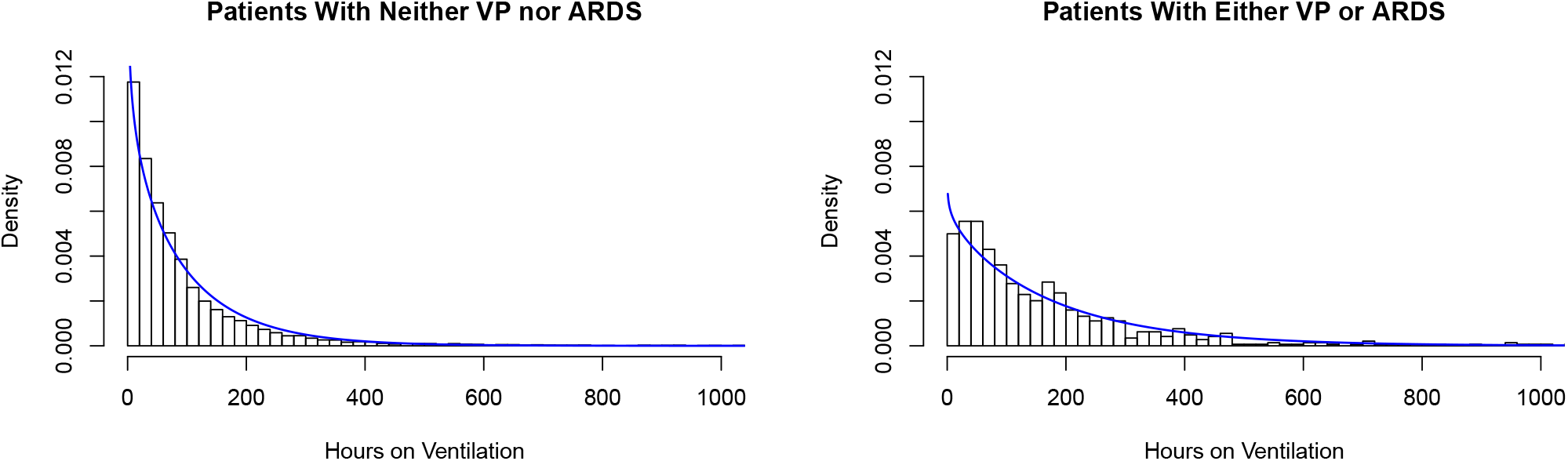
Density histograms of the time on mechanical ventilation for two patient groups. Fitted gamma distributions are overlaid as a blue curve.

Figure 4 displays the 2017–2018 monthly proportion of ventilation starts that are for patients with either VP or ARDS, as opposed to patients with neither diagnosis. Both years show a seasonal trend of higher winter proportions of patients with VP or ARDS, compared to the summer months. We modeled the arrival rates for these patients using a time-dependent mixing probability based on the monthly proportion of ventilation starts for patients in this group in both 2017–2018, which is also displayed in Figure 4. The compliment of this time-dependent mixing probability gives the arrivals for the group of patients with neither VP nor ARDS.

**Fig. 4.**
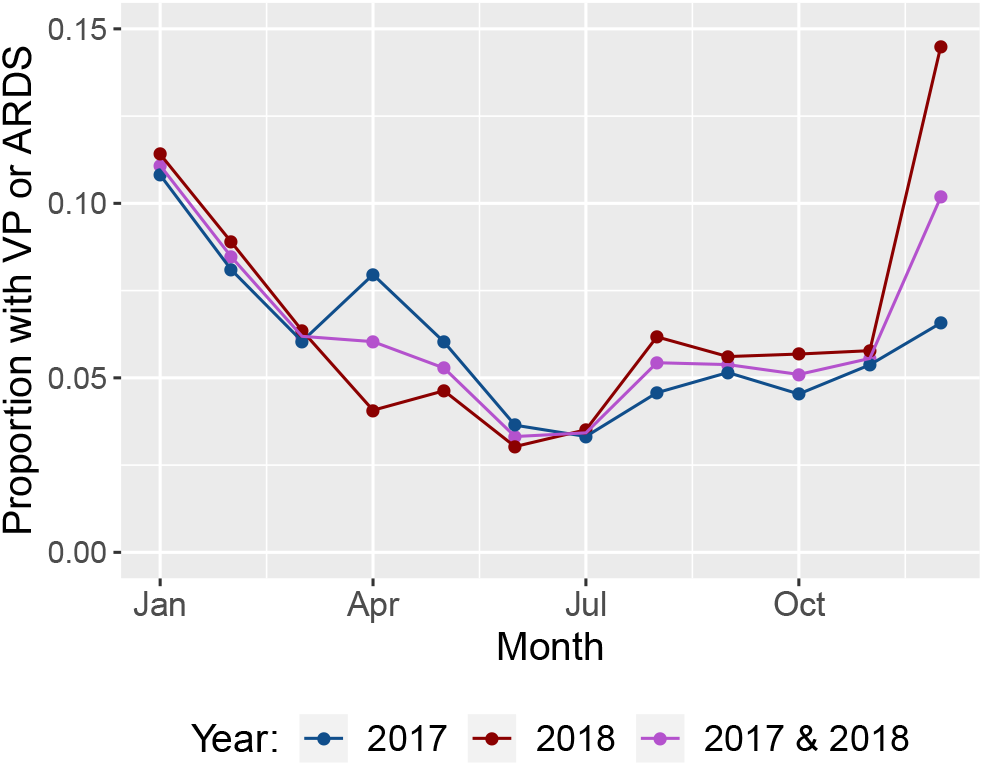
Monthly proportion of ventilation starts for patients with either VP or ARDS. Values are shown for 2017, 2018, and both years.

Symptom onset time is not routinely entered in medical record data in BC, and we are unaware of BC specific estimates for the distribution of time between COVID-19 symptom onset and severe symptoms requiring mechanical ventilation. A study by Phua et al. [51] estimated the median time from symptom onset to ICU admission as 7–12 days. Therefore, we assumed a uniform distribution between 7 and 12 days for our symptom delay model.

### 5.3 COVID-19 Case Projections

The BC Centre for Disease Control provided COVID-19 case projections made using a stochastic disease model^1^ based on Hellewell et al. [29]. Their epidemiological model was calibrated using historical data on cases in BC and projects cases under different scenarios by varying a transmission rate parameter. Scenarios of reduced transmission were specifically used to illustrate the impact of public health measures such as social distancing and other changes in population behavior. Significant public health measures in BC during the first wave of the epidemic began on March 16^th^ and included restricting gatherings to no more than 50 people and closing schools^2^. Their model was re-calibrated weekly in March and April 2020, with projections extending one month from calibration date. The results in our paper are based on projections released on March 19^th^, 2020 for COVID-19 case counts from March 16^th^ to April 13^th^.

Based on expert opinion and initial data on the epidemic in BC, we assumed that 6.7% of COVID-19 cases would require ICU care. We further assumed that 70% of COVID-19 ICU patients would require mechanical ventilation, based on the ICNARC Report on COVID-19 (April 10^th^, 2020) [30] and expert opinion on hospitalizations in BC. We multiplied the BC CDC projected daily case counts by the above two proportions, in order to project COVID-19 symptom onset for cases which will eventually require mechanical ventilation. We then converted this into projected hourly rates at noon of each day, and linearly interpolated to obtain instantaneous rates for the symptom delay model.

## 6 Results

We present the results of applying the multi-class Erlang loss model from Section 3 to project and optimize ventilator capacity at a provincial level in BC, during the first wave of the COVID-19 epidemic in March and April 2020. Validation of the model for historical non-COVID-19 ventilator use is presented in Subection 6.1. Model projections of ventilator access are presented in Subsection 6.2, under different epidemic scenarios to illustrate the impact of public health measures. Subsection 6.3 compares access projections from simulation with results from three numerical approximation methods: PSA, MOL, and FPA. Results of our hybrid capacity optimization approach are presented in Subsection 6.4.

### 6.1 Validation

We validated our non-COVID-19 modeling assumptions by comparing simulated non-COVID-19 ventilator use with pre-COVID-19 ICU data. Using our BC ICU Database extract, we estimated the number of patients on a ventilator at any time from the recorded start and stop times of ventilation. For the 10.4% of records without a recorded stop time, we assumed a stop time proxy equal to the start time plus an annual mean ventilation time. This yielded a time series of estimated ventilator utilization for 2017 and 2018.

For comparison with historic data, we ran our DES implementation with only non-COVID-19 ventilator demand for the 20 hospitals in the BC ICU Database, and without a reduction in elective surgeries. We used a ventilator capacity of 356, which corresponds to the estimated number of functional adult ventilators in these hospitals. We ran the DES model for a three-year simulation of 2016 through 2018. We measured the mean, 5^th^, and 95^th^ percentiles of the number of simulation ventilators in use, which is compared in Figure 5 with the inferred historic ventilator use for 2017 and 2018. Simulation output for 2016 was not analysed, because the first simulation year was used to populate the model. In Figure 5, the arrival rates and patient group proportions are given by monthly averages across data from 2017 and 2018. With these monthly arrival rates, the 5^th^ to 95^th^ percentile range of the simulation output covers the historic data 84.5% of the time. This indicates some unaddressed data variability; however, a monthly time granularity is appropriate for the purpose of projecting non-COVID-19 ventilator demand into 2019–2020.

**Fig. 5.**
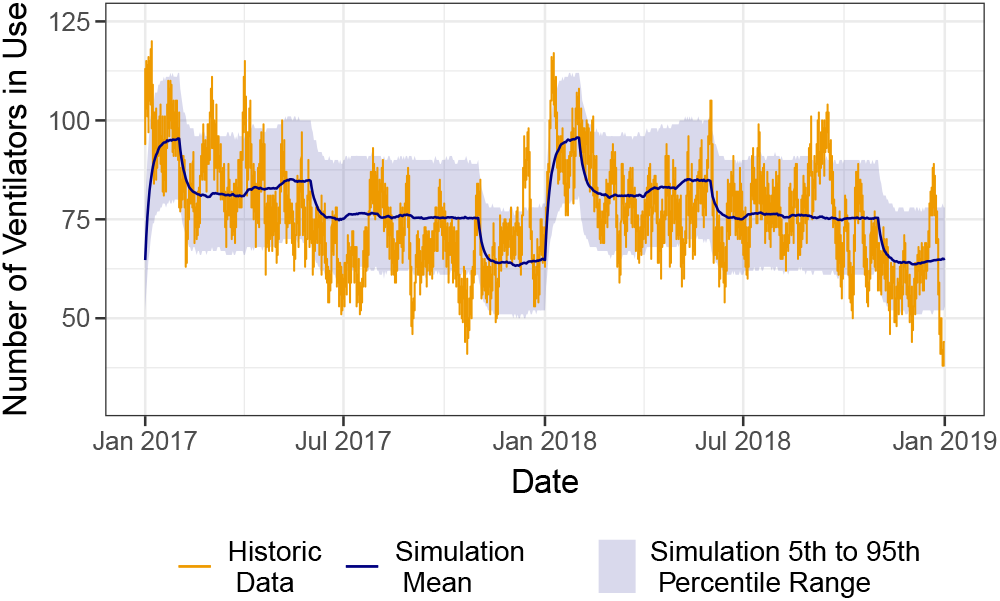
Comparison of pre-COVID-19 simulation and inferred 2017-2018 ventilator use from the ICU DB. This simulation uses monthly average arrival rates over 2017 and 2018. The mean, 5^th^, and 95^th^ percentiles of simulation ventilator use are measured from 4000 runs, using 2016 to populate the model.

We performed a second simulation evaluation with increased time granularity in the arrival rate to further validate our model against historical data. Figure 6 shows these simulation results, in which arrival rates are specific to each year and week of the simulation, and patient group proportions are given by year and month. With this level of time granularity, the simulation is able to fully match the data variability, since the 5^th^ to 95^th^ percentile range covers the historic data 90% of the time. Since the unmet variability in Figure 5 can be fully addressed by increased time-granularity of arrivals, it does not indicate limitations in our other non-COVID-19 modeling assumptions.

**Fig. 6.**
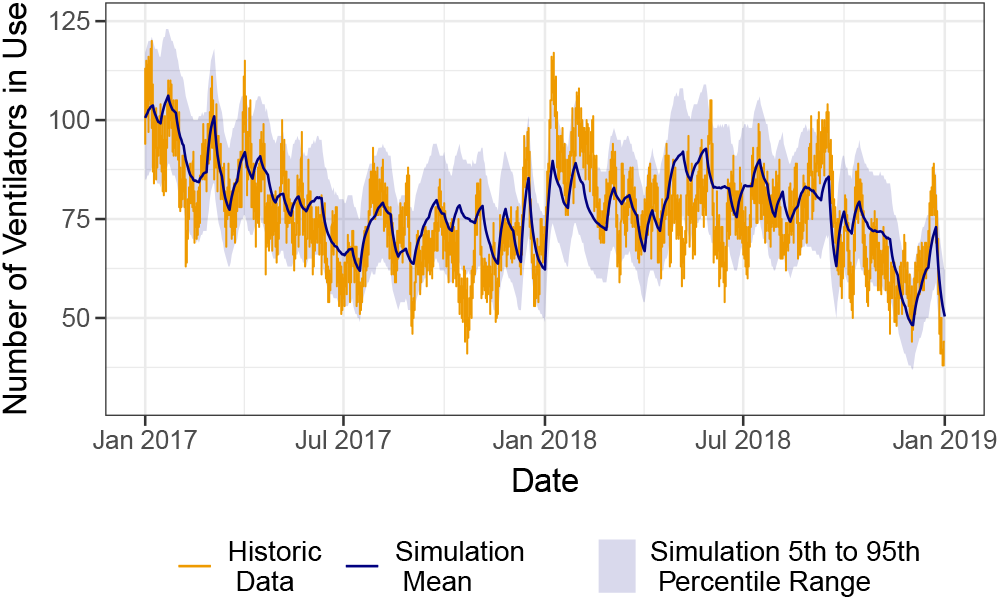
Comparison of pre-COVID-19 simulation and inferred 2017-2018 ventilator use from the ICU DB. This simulation uses week and year specific arrival rates for 2016, 2017 and 2018. The mean, 5^th^, and 95^th^ percentiles of simulation ventilator use are measured from 4000 runs, using 2016 to populate the model.

### 6.2 Projected Ventilator Access

We simulated the queue model using DES to project provincial ventilator access under different epidemic trajectories, during the first wave of COVID-19 in BC. Model simulation results for the March 19^th^, 2020 case projections from the BC CDC are shown in Figure 7, under scenarios with and without reduced transmission due to public health measures. These projections start at the beginning of the community spread of COVID-19 on March 16^th^, 2020, and project COVID-19 case counts until April 13^th^, 2020. To build up non-COVID-19 ICU occupancy, we started the simulation one year prior to the epidemic projections. We ran the simulation until April 20^th^, 2020, since the delay between symptom onset and ventilation has a minimum of 7 days.

**Fig. 7.**
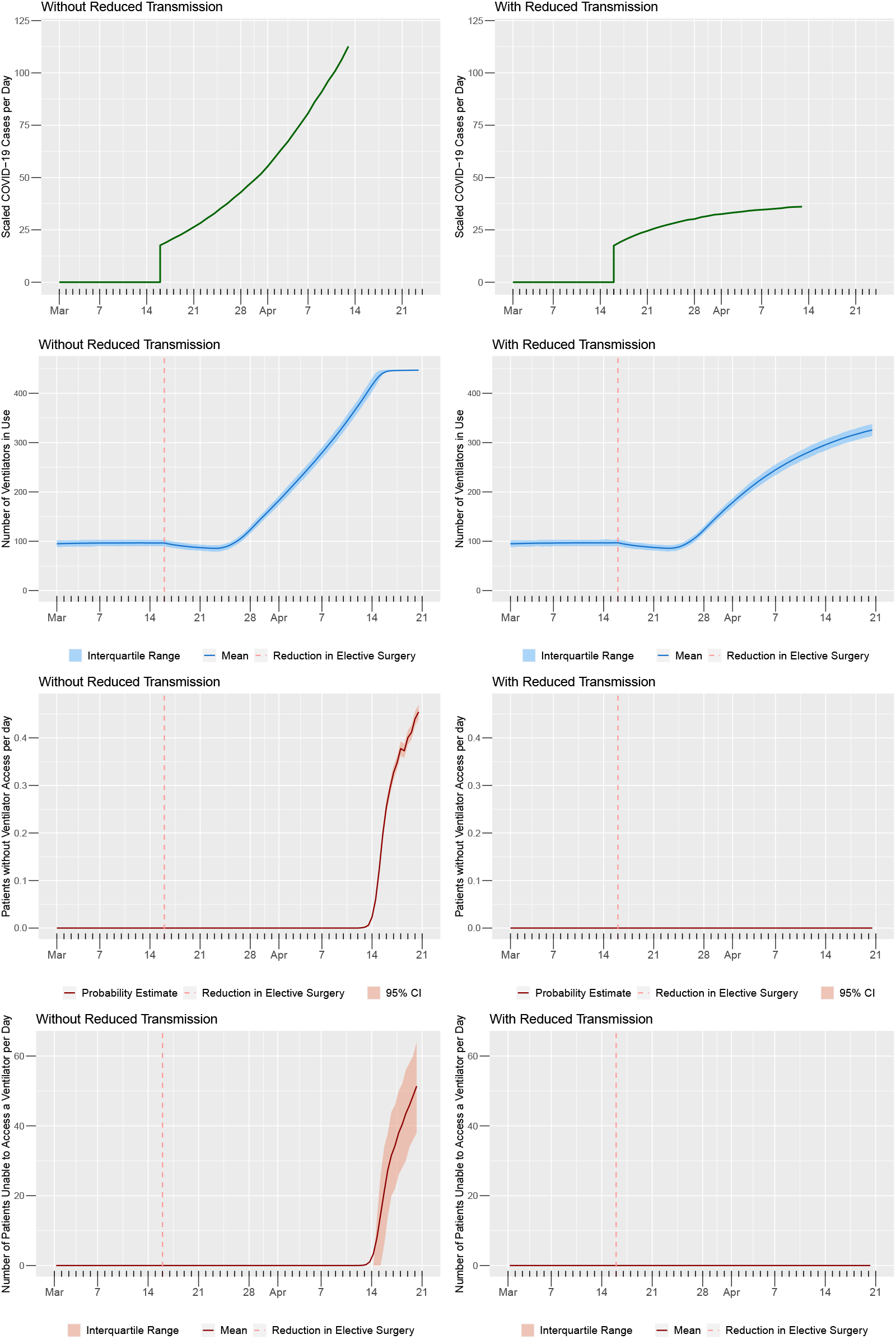
Epidemic projections and DES results for scenarios with and without reduced transmission. Simulation runs use 448 ventilators. The dotted red line represents the start of projections and the reduction in elective surgeries. The topmost panels present epidemic projections of daily COVID-19 cases requiring a ventilator, from March 16^th^ to April 13^th^, 2020. The second and third pairs of panels shows DES estimates for the expected number of ventilators in use and loss probability, both measured at midnight and noon of each simulated day. The fourth pair of panels show the average number of patients unable to access a ventilator per day, measured every half-day. Simulation results are obtained using 4000 runs.

The effect of the cancellation of non-urgent elective surgeries on March 16^th^ is noticeable in both scenarios as a slight decrease in ventilator utilization, before an increase occurs due to the projected rising COVID-19 cases. Without reduced transmission, the projected number of COVID-19 cases requiring a ventilator reaches approximately 112 patients per day by the end of the projection. In this scenario, the estimated probability of reaching ventilator capacity is negligible until approximately April 14^th^, when it begins to rise dramatically. By the end of the simulation, the mean number of patients unable to access a ventilator reaches approximately 50 per day. However, with reduced transmission due to public health measures, the projected rate of COVID-19 ventilator cases reaches a substantially lower rate of approximately 36 patients per day. In this scenario, the estimated probability of reaching ventilator capacity remains negligible and all the simulated patients are able to access a ventilator.

### 6.3 Comparing Numerical Approximations

We applied three numerical methods, namely PSA, MOL, and FPA, to approximate time-dependent ventilator utilization and access under the epidemic scenario without reduced transmission from the March 19^th^ epidemic projections. Figures 8 and 9 compare the results of these methods with simulation values for the expected number of ventilators in use and loss probability. The percentages of time that these estimates are within the DES interquartile range (IQR) or confidence interval (CI) are shown in Table 2, for March 16^th^ onwards. PSA, MOL, FPA, and simulation values were all evaluated at 12-hour intervals aligned with noon and midnight of each day. All of the integration in the numerical methods was performed using the trapezoid rule. Simulation estimates were obtained using 4000 runs and a tolerance value of 10^*−*10^ was used to obtain FPA results. Table 3 compares peak loss probability estimates, as well as computation times for these four approaches.

**Fig. 8.**
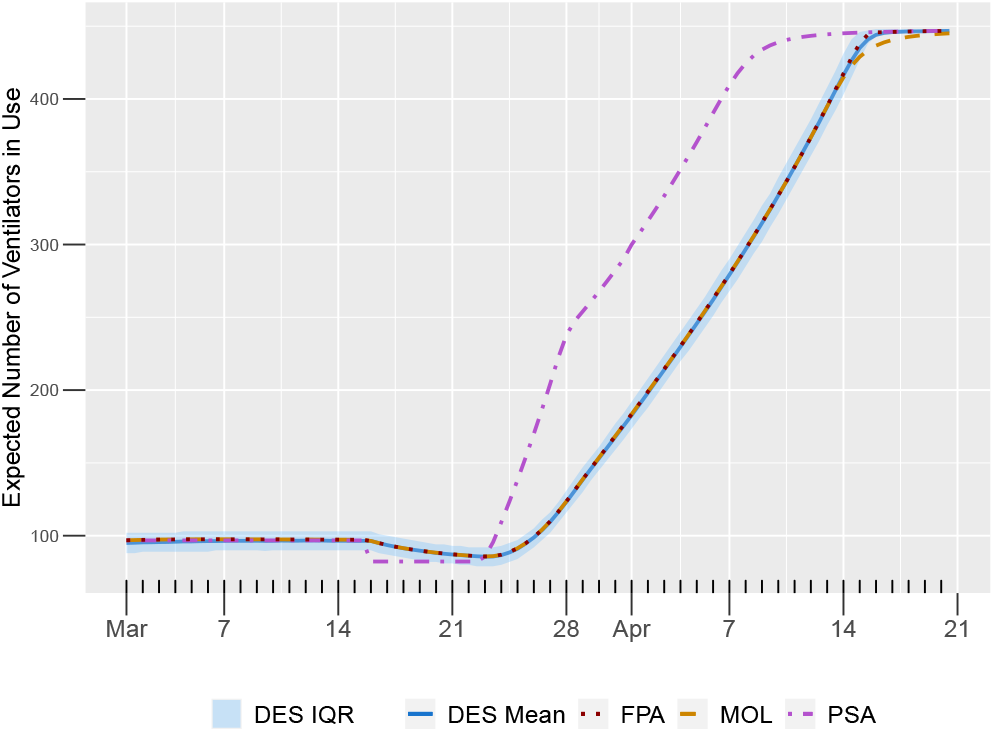
Comparison of four estimates for the time-dependent expected number of ventilators in use. These projections use the current ventilator capacity of 448, and assume an an epidemic scenario without reduced transmission. The mean and interquartile range (IQR) for simulation values are obtained using 4000 runs.

**Fig. 9.**
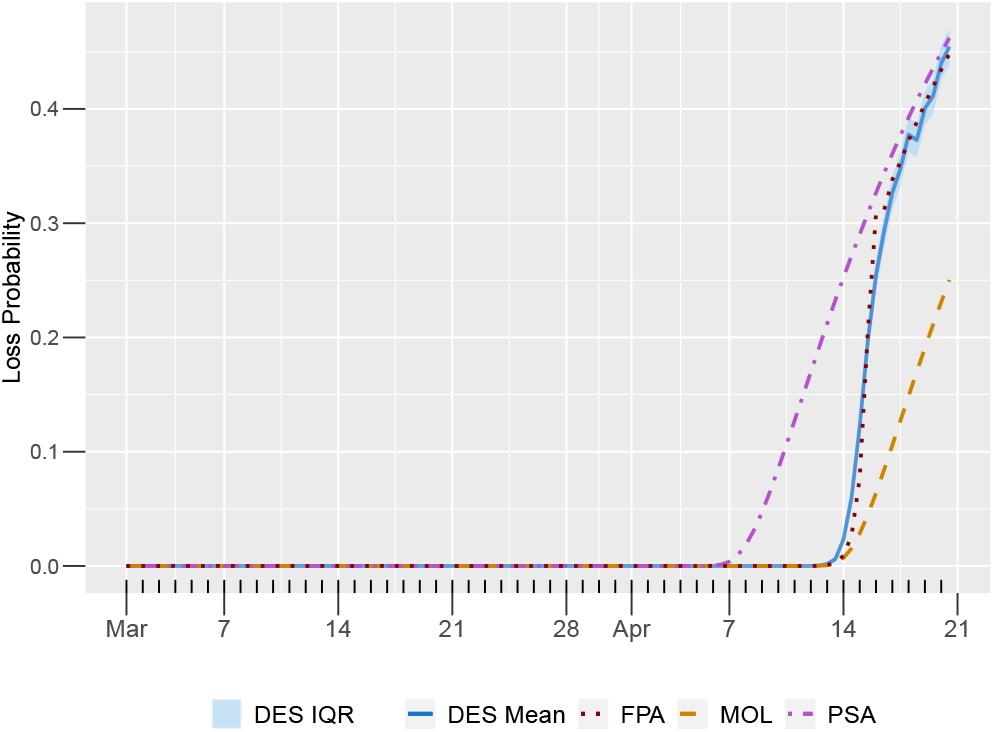
Comparison of four estimates for the time-dependent loss probability. These projections use the current ventilator capacity of 448, and assume an an epidemic scenario without reduced transmission. The mean and interquartile range (IQR) for simulation values are obtained using 4000 runs.

**Table 2.** Comparison of the percentage of time-dependent estimates that are within the simulation ranges, for twice daily estimates between March 16^th^ to April 21^st^, 2020. All of the estimates are under the epidemic scenario without reduced transmission and use the current supply of 448 ventilators. The simulation range for the expected number of ventilators in use 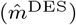 is an interquatile range, and the range for the loss probability 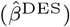 is a 95% confidence interval.

| Method | Percentage of Time within DES Range for |  |
| --- | --- | --- |
| | $\hat{m}^{\text{DES}}$ | $\hat{\beta}^{\text{DES}}$ |
| PSA | 27.8% | 61.1% |
| MOL | 84.7% | 79.2% |
| FPA | 100% | 90.2% |

**Table 3.** Peak estimated loss probability for DES, PSA, MOL, and FPA methods, as well as computational times. Computation times were evaluated on a 2.3 GHz quadcore Intel core i7 processor. DES results were obtained from 4000 runs and were executed in parallel over 8 cores.

| Method | Peak loss probability (95% CI) | Evaluation time |
| --- | --- | --- |
| DES | 0.455 (0.44,0.47) | $\geq 25$ m |
| PSA | 0.462 | 0.34 s |
| MOL | 0.250 | 1.06 s |
| FPA | 0.448 | 2.08 s |

The FPA estimates are substantially closer to the simulation results than estimates from PSA and MOL. FPA values for the expected number of ventilators in use are within the simulation IQR 100% of the time, and values for loss probability are within the simulation CI 90.2% of the time (Table 2). Both FPA and PSA estimates capture the peak loss probability (Table 3), but PSA results predict that capacity would have been reached about a week sooner than in the simulation (Figure 9). MOL substantially underestimates the peak loss probability, yielding an estimate which is 45% less than the simulation (Table 3). In our implementations, FPA requires almost double the computational time of MOL; however, FPA is still orders of magnitude faster than DES (Table 3).

### 6.4 Capacity Optimization Results

We performed the search procedures described in Subsection 4.2 to determine the ventilator capacity required to keep ventilator access above 95% in the March 19^th^ epidemic scenario without reduced transmission. This section compares proximate capacity requirements based on the three numerical approximations (PSA, MOL, and FPA) to DES optimization results obtained using a hybrid approach.

First, we performed three separate linear searches using the PSA, MOL, and FPA model approximations. Table 4 compares the proximate required ventilator capacity for each of these methods to the DES-based result. Our hybrid approach to simulation-based optimization initializes an iterative RSM search procedure with the FPA proximate capacity requirement. This starting point was informed by the results in Subsection 6.3, which demonstrate that FPA performs the best at predicting loss probability under the current ventilator capacity.

**Table 4.**
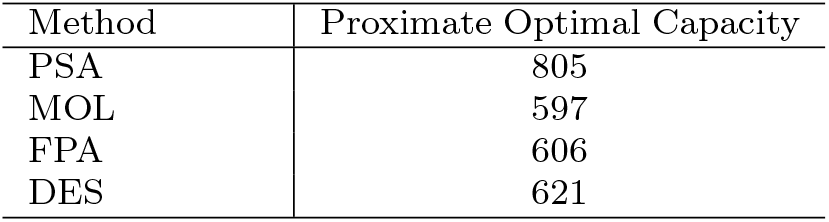
Ventilator capacity requirements to keep the respective loss probability estimates (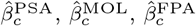, and 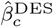) under 5%.

| Method | Proximate Optimal Capacity |
| --- | --- |
| PSA | 805 |
| MOL | 597 |
| FPA | 606 |
| DES | 621 |

Our simulation-based optimization started at the FPA proximate capacity requirement of 606 ventilators, and took only 3 iterations of second order RSM approximations until consecutive estimates were within 5 ventilators of each other. Figure 10 presents the DES loss probabilities for the 10 values surrounding the last RSM capacity estimate, and it shows at least 621 ventilators are required to maintain a 5% target. A reduction in capacity of up to 5 ventilators will keep the loss probability within 6%.

**Fig. 10.**
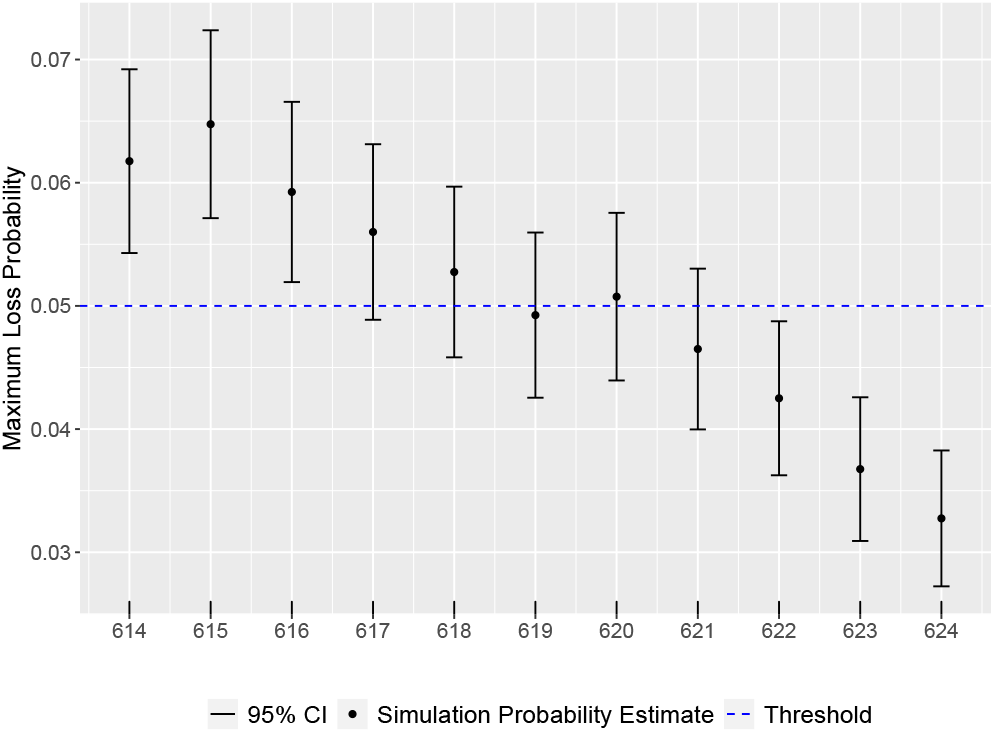
Linear search across the ten ventilator capacities surrounding the final RSM required capacity estimate. For each number of ventilators, the peak loss probability and 95% confidence interval (CI) is estimated using 4000 simulation runs.

For the optimal capacity of 621 ventilators, Figures 11 and 12 compare DES and numerical approximation projections of ventilator use and access. In these figures, MOL and FPA both accurately approximate the expected number of ventilators in use. However, both FPA and MOL methods underestimate loss probability.

**Fig. 11.**
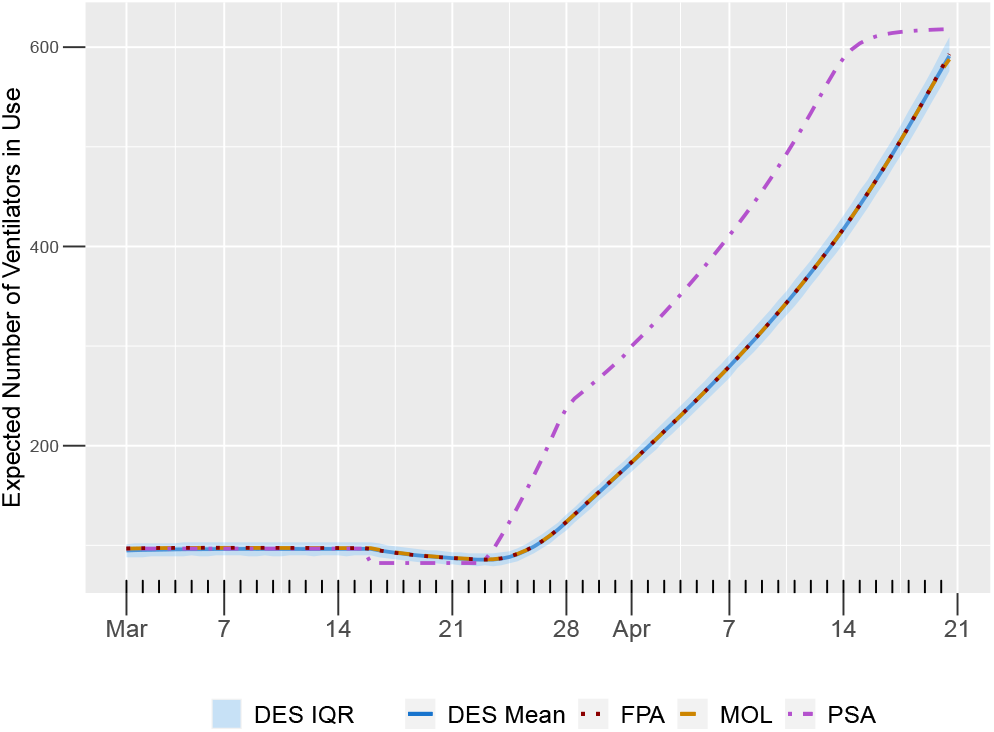
Comparison of the expected number of ventilators in use, for the three numerical approximation methods (PSA, MOL, and FPA), with an optimal capacity of 621 ventilators under the epidemic scenario without reduced transmission. Simulation mean and interquartile range (IQR) values are obtained from 4000 runs.

**Fig. 12.**
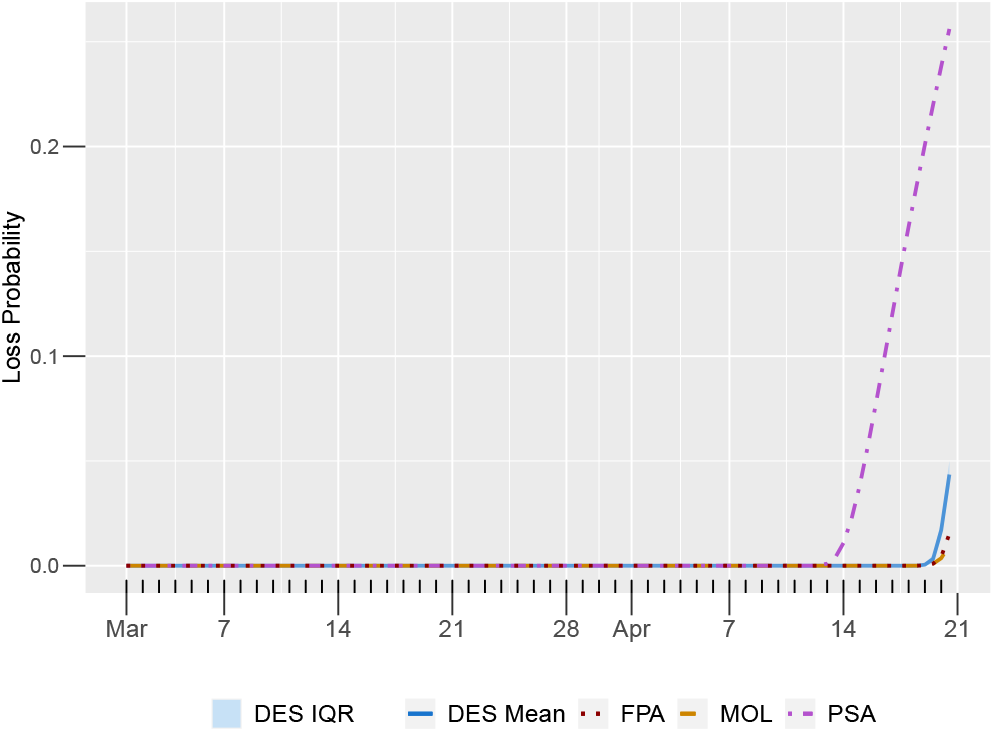
Comparison of estimated loss probability, for the three numerical approximation methods (PSA, MOL, and FPA), with an optimal capacity of 621 ventilators under the epidemic scenario without reduced transmission. Simulation mean and 95% confidence interval (CI) values are obtained from 4000 runs.

## 7 Discussion and Conclusions

We applied a multi-class Erlang loss model to inform ventilator capacity management during the first wave of the COVID-19 pandemic in BC, Canada. We worked closely with analysts at the BC Ministry of Health, the Provincial Health Services Authority, and the BC Centre for Disease Control in March and April 2020, to provide weekly reports on projected ventilator access. We collaborated with these organizations to obtain up-to-date BC specific data and parameter values for our model, including both non-COVID-19 critical care use and weekly updated COVID-19 case projections. The results presented in this paper are based on the March 19^th^ epidemic projections from the BC CDC, under different levels of transmission. Simulation results predict that under a scenario of reduced transmission of COVID-19—through means such as social distancing, but also other public health measures and changes in population behavior—ventilator capacity would likely not be reached, thereby helping to avert as many as 50 deaths per day within the time frame of March 16^th^ to April 20^th^, 2020. Under the COVID-19 projections without reduced transmission, an additional 173 ventilators would have been required to ensure that the probability of immediate patient access to a ventilator is at least 95%. However, with public health interventions including social distancing, the current ventilator supply was sufficient. These results were provided to health system analysts and policy makers to help inform capacity management decisions, including whether to expand the current ventilator supply. Additionally, our model links public health measures to operational impacts on ventilator access. Our work provides a tool for policy makers to quantify the interplay between public health interventions and critical care access.

Our projections of ventilator access are dependent on the underlying epidemic projections and need to be interpreted in the light of the challenges with predicting the epidemic trajectory [35, 61]. In particular, our results are highly sensitive to estimates of the peak in the epidemic wave. Furthermore, public health measures are often implemented in response to epidemic projections and these measures may change the course of the epidemic.

Another contribution of this study is the comparison of PSA, MOL, and FPA under rapidly changing epidemic demand. Previous studies have focused on evaluating these numerical methods under sinusoidal demand [3, 16, 19, 21, 22, 24, 25, 23, 32, 34]. Our analysis compares the accuracy and efficiency of these numerical approximation methods for projecting ventilator access under the rapid growth in the March 19^th^ COVID-19 projections without reduced transmission. With the current ventilator supply, FPA results closely approximate the simulated loss probability throughout the duration of the projection. In comparison, MOL results underestimate the peak loss probability within this time frame and PSA results overrespond, predicting capacity would be reached about a week early. The FPA estimate of the number of ventilators required to meet access targets is closer to the simulation capacity requirement than the PSA and MOL estimates. However, FPA underestimates capacity requirements by 15 ventilators (2.5%). The comparative accuracy of FPA is manifest, because it improves MOL estimates by iterating to capture the time-dependent interaction between capacity, access, and utilization [3, 32]. However, FPA updates still rely on steady-state formulae, and its accuracy is not guaranteed for highly time-dependent systems. Our results demonstrate the extent of the accuracy of FPA under epidemic-type demand. Although FPA requires more computational time than PSA and MOL, it is still substantially faster than DES.

Epidemic projections are frequently updated during the COVID-19 pandemic, and efficient model analysis is important for timely support of decision making. We developed a hybrid capacity optimization search by drawing on our comparison of numerical approximations. Since FPA was the most accurate for projecting ventilator access under the current capacity, we used FPA to find a proximate initial value for an iterative simulation-based capacity search. By starting at this point, the optimal capacity range was identified in only three iterations of a modified RSM search. Our hybrid approach combines the accuracy of DES with boosted efficiency from FPA, which addresses the computational challenges of loss model capacity optimization under rapidly growing epidemic demand.

Overall, this study provides valuable insight for current and future epidemic capacity planning. Our comparison of numerical approximation methods motivates the further use of FPA in epidemic queue modeling and capacity optimization. Our hybrid search procedure addresses the computational challenges of optimizing loss models under rapidly growing demand. This enables further application of loss models to inform epidemic capacity planning in the context of patient-centred access indicators.

As the COVID-19 pandemic progresses, our analysis can be further updated using the latest case projections, parameters for ventilator use, and treatment protocols. By using case projections from epidemic models that incorporate vaccination, our model could link vaccine uptake to the utilization of critical care resources. The queue model can be adapted to other geographic regions by incorporating location-specific critical care data. Future work could consider modeling additional medical resources, for example ICU beds and respiratory therapists, and staff scheduling could be informed by increasing the time granularity of capacity optimization. More sophisticated optimization techniques could be incorporated to provide greater computational efficiency to address these extensions. Our framework is widely applicable to many critical care resources that can face surge demand.

## Data Availability

A github reference is given in the manuscript for BC Centre for Disease Control case projections used in the model.

https://github.com/bcgov/epi.branch.sim

## Acknowledgements

We are grateful to Sandra Feltham and the Hospital & Diagnostics Analytics Team at the British Columbia (BC) Ministry of Health for supporting this project and providing summary statistics from the Discharge Abstract Database. We thank the epidemiological modeling team led by Michael Otterstater at the BC Centre for Disease Control for providing weekly COVID-19 case projections from their model; Ognjenka Djurdjev at the Provincial Health Service Authority for data on the number of ventilators available; Lena Farina at St. Paul’s Hospital, Vancouver, for expert opinion on ventilator maintenance and cleaning times; and Donald Griesdale at Vancouver General Hospital, for insight into critical care operations.

## Appendix A Response Surface Methodology

We used a modified response surface methodology (RSM) procedure to solve the capacity optimization problem in equation (20), which identifies the minimum ventilator capacity required to meet at target on the DES loss probability. Our approach is based on the RSM framework in Nicolai and Dekker [47]; however, we made several modifications based on our problem context. This appendix describes the algorithmic details of our implementation; higher level discussion is in Subsection 4.2.

In our RSM application, the response function is the maximum simulation loss probability as a function of the number of ventilators *c*. We denote the iterative estimates of the required ventilator capacity by *c*_*i*_ and the iterative radii of the successive regions of interest by *r*_*i*_, for *i* = 0, 1, 2, … We begin with initial estimated capacity *c*_0_ and radius *r*_0_ = round(0.1 *c*_0_). The target loss probability is denoted by *α*. The details of our implementation are as follows:

Iterate the following over *i* = 0, 1, 2, …, until *r*_*i*_ *≤* 5 or |*c*_*i*_ *− c*_*i−*1_| *≤* 5:

a. Use DES to estimate the maximum loss probability at each ventilator capacity value in a one dimensional central composite experimental design [47]. This design involves five estimates made at the center value *c*_*i*_, and one estimate made at each of the values: *c*_*i*_ *− r*_*i*_, round(*c*_*i*_ *−* 0.5 *r*_*i*_), round(*c*_*i*_ + 0.5 *r*_*i*_), and *c*_*i*_ + *r*_*i*_. For each capacity value, the maximum loss probability is estimated using 200 simulation runs.
b. Using these estimated points, perform a least-squares regression fit to a second-order model,

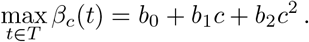
c. If the estimated coefficients satisfy 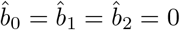, then set *r*_*i*+1_ = 2 *r*_*i*_ and move to the next iteration. This check increases the region of interest if the DES estimate of the response function is zero. Note that the response function is asymptotic to zero for a large number of ventilators.
d. If the overall regression fit is statistically significant (F-statistic has p-value at most 0.05), then set

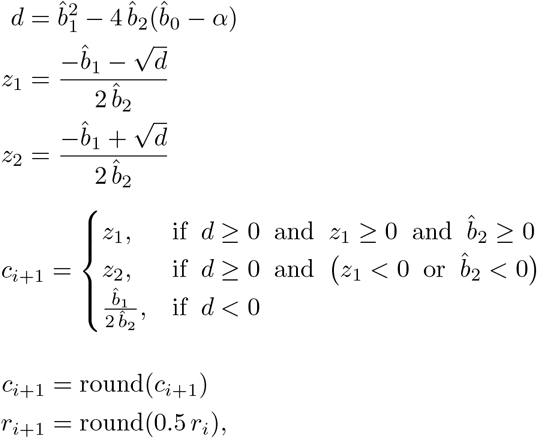

and move to the next iteration.
e. If the overall regression fit is not statistically significant (F-statistic has p-value greater than 0.05), then double the number of runs per DES evaluation, until all are statistically significant.

## Footnotes

1 The BC CDC model is implemented as an R package, which is available from https://github.com/bcgov/epi.branch.sim.

2 Additional details on public health measures undertaken in BC are available at https://news.gov.bc.ca/releases/2020HLTH0086-000499.

